# Face Mask Fit Hacks: Improving the Fit of KN95 Masks and Surgical Masks with Fit Alteration Techniques

**DOI:** 10.1101/2020.10.28.20221895

**Authors:** Eugenia O’Kelly, Anmol Arora, Sophia Pirog, Charlotte Pearson, James Ward, P John Clarkson

## Abstract

The COVID-19 pandemic has drawn unprecedented attention to the use of masks and fabric face coverings to prevent the spread of respiratory viruses, such as SARS-CoV-2. The fit of a mask has been identified as one the primary factors in determining the effectiveness of masks. If substantial gaps exist between the mask and the wearers face, air may take the path of least resistance through gaps and avoid filtration, both during inhalation and exhalation. A number of techniques, referred to as fit hacks have emerged to improve the fit of face masks. In this paper we test a variety of fit hacks on surgical masks and KN95 masks to compare their effectiveness. We identify fit hacks which greatly improved the fit of masks, and thus their effectiveness.

## Introduction

During the COVID-19 pandemic, shortages of personal protective equipment (PPE) have resulted in members of the public having no choice other than to wear poorly fitting face masks, such as surgical and KN95 masks, which are the focus of this study. Proper fit has been noted as a primary factor in determining the effectiveness of face masks *(1)* but the masks available to the public are often accompanied by issues of poor fit. In an effort to improve the protection such masks offer, many people have designed alterations, a.k.a. fit hacks, in an attempt to improve fit. While these fit hacks have garnered widespread attention and media coverage, the impact of most hacks on fit remain untested. Such fit hacks include the use of pantyhose over the mask *(2)* and altering the mask shape by knotting the ear bands *(3)*.

Better fitting masks offer fewer gaps between the wearer’s face and the edges of the mask. Decreasing these gaps, and thus increasing fit, improves not only the protection the mask can offer the wearer from airborne particles but also offers greater protection to the public. In addition, better fit for the wearer has been theorized to reduce the degree of SARS-CoV-2 inoculum, thus potentially eliminating or reducing the severity of an infection *(4)*.

Unfortunately, little research has been done to assess the effectiveness of these techniques. Early research, seeking to validate a protocol for measuring mask performance, has suggested that a nylon over-layer worn over a face covering may enhance the performance of masks. However, the same data also suggests that improvement techniques may heterogeneously affect different types of face coverings *(2)*. This remains an understudied area of research with a notable shortage of multi-participant studies.

This study aims to answer the research question: “Do simple fit hacks actually improve the fit of face masks?” Our study explores the quantitative fit score of several masks with and without a range of fit hacks applied. In doing so, this research seeks to discover which, if any, techniques may be used to improve the fit of face masks aimed primarily for use by the general public. Seven fit hacks were tested. Photos of the applied fit hacks can be found in Fig. 1.

**Fig. 1:**
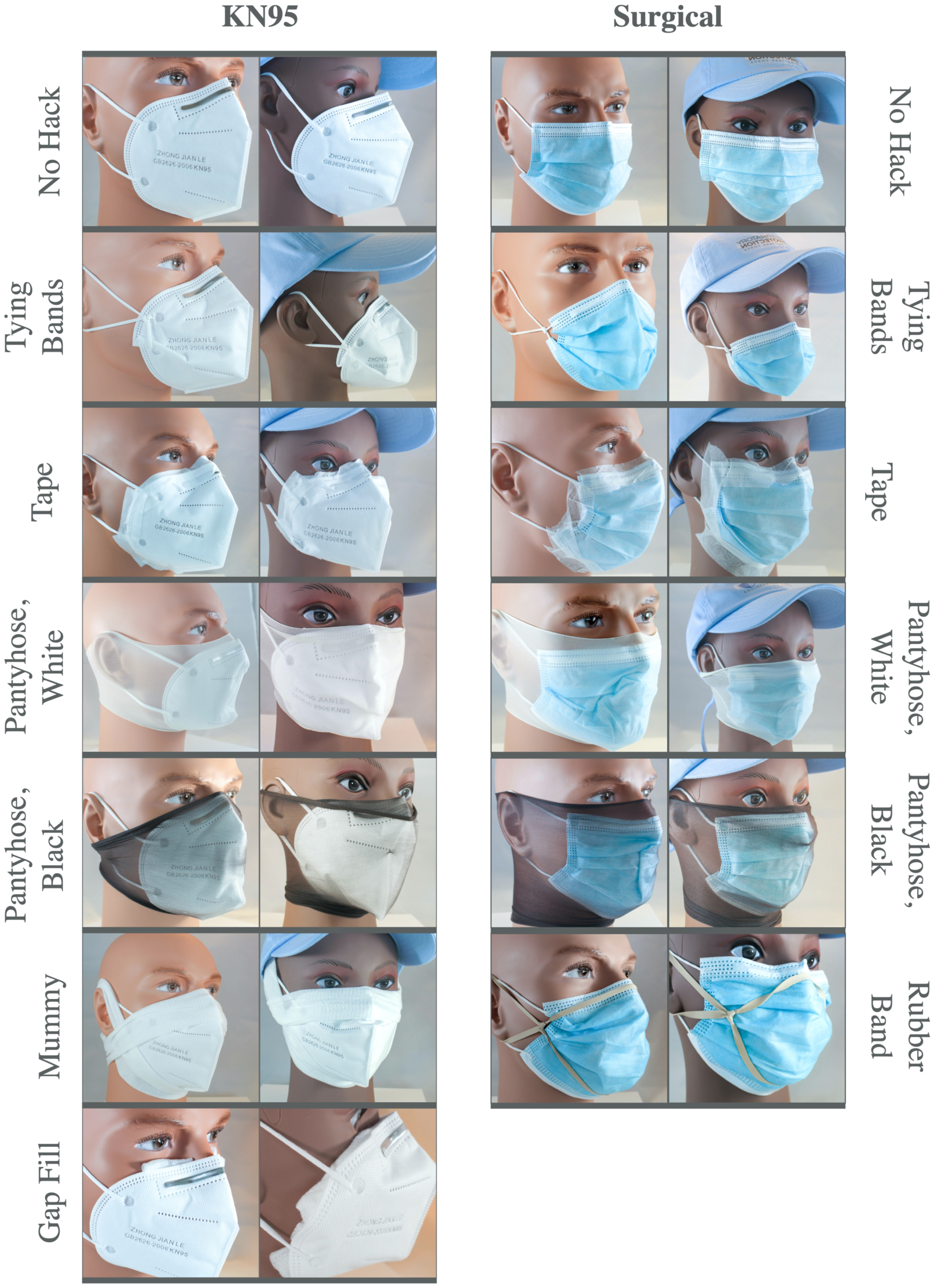
Pictures of the tested fit hacks applied to KN95 and surgical masks on two mannequin heads. The male mannequin head is the size of an average male, while the female represents the size of a small female head or young teen head.

## Materials and Methods

There are two established methods used to assess the fit of face masks: quantitative fit testing and qualitative fit testing. Quantitative fit testing is the most robust and accurate method, providing a nuanced measurement of degree of fit. As such, quantitative fit testing was utilized in this study to evaluate the efficacy of fit hacks.

In quantitative fit testing, a particle counter continuously measures the concentrations of particles both inside and outside of a mask while worn. The concentration of particles outside the mask are divided by the concentration of particles inside the mask, via an industry standard formula, to generate a fit factor *(5,6)*. This fit factor is a numerical score of how well the mask fits the wearer; meaning, the fewer particles that make their way into the mask, it is assumed to be a better fit. For masks with less than 99% efficiency, fit factor scores can range from 1 to over 200 (represented as 200+). Higher scores indicate better fit while low scores indicate a poor fit. When the fit of N95 or FFP3 masks are assessed, a score of at least 100 is required for the mask to be considered to have adequate fit *(7)*.

For this study, quantitative fit testing was performed with a Portacount, TSI, Minnesota, Model 8038+ according to OSHA protocol 29CFR1910.134. Particles ranging from 0.02 micrometers to over 1 micrometer were measured. Accuracy using the 8038 is typically +/- 10% of a given reading. A particle generator from TSI, Minnesota, Model 8026 was used to atomize sodium chloride (NaCl) particles during testing.

Four participants took part in the study, three female and one male. Participants completed seven activities, intended to reproduce a range of occupational activity, according to OSHA protocol 29CFR1910.134. These activities include normal breathing, deep breathing, talking, bending over, and turning the head. Due to the length of the study, over two hours per participant, participants were allowed to sit during certain sections of the test.

A range of fit hacks, taken from the internet and from observing the public, were tested with two masks: a KN95 mask and a surgical mask. First, fit scores of the mask without a fit hack were compared with the fit scores once the fit hack was applied in order to determine the fit hack’s impact.

In this study, we focus on KN95 and surgical masks as both masks are widely available and, if fit properly, can offer a high degree of filtration. KN95 masks promise similar filtration benefits to N95 masks; however, unlike N95 masks, the public’s access to KN95 masks has not been restricted by this COVID-19 pandemic. While KN95 masks offer significant filtration potential, their poor fit on many individuals negates the potential benefit of these masks *(8)*. Surgical masks are in common use and, if constructed out of the proper materials, can offer a very high filtration ability.

Seven fit hacks were tested, as shown in Fig. 1 and as described below:

Tape: The edges of the mask are sealed with cloth tape. Care was taken to mold the tape to the face and to seal any gaps between the skin and the edge of the mask.

Stuffed Gaps: First aid gauze was used to fill visible gaps in the mask, until no visible space between skin and mask remained.

Mummy: A roll of first-aid gauze was used to tightly bind the mask to the face, pressing the edges of the mask to the skin of the face.

Panythose: Two brands of pantyhose were placed over the head to press the mask into place, a method first proposed and tested by Mueller et al *(2)*.

Knotting Ear Loops: To make a large mask fit a smaller face, an overhand knot was made of the ear loop elastic near the mask. This hack gained media attention when dentist Dr. Olivia Cui posted a video of herself performing the hack on TikTok *(3)*.

Rubber Bands: In a hack proposed by Apple engineers, three rubber bands are used to create a ‘brace’ *(9)*.

Four participants took place to determine if the benefits incurred by the application of a fit hack differed significantly between individuals. Participants are identified here by their respective gender and age. Participant F-29 had the smallest head size with a circumference of 54cm. Participants F-51 and F-18 had a circumference of 55cm and 56cm respectively. M-20 had the largest head size, with a circumference of 60cm.

Our study should not be taken as an endorsement of any method nor as a definitive indication of which hacks may work best. While our results may suggest some techniques are more effective than others at improving fit, facial features are likely to alter the degree of improvement at an individual level.

All hacks were tested with two exceptions. One participant was unable to fit the KN95 mask with ear bands tied, as the adjustment caused the wire of the mask to rub against a sore. Another participant was unable to fit pantyhose Brand A over his head. In these cases, the hack was not tested.

## Results

As shown in Fig. 2, the pantyhose and tape hacks were most effective at improving fit in KN95 masks, although significant variation between participants occurred. The use of pantyhose produced improvement in the fit of KN95 masks depending on brand, with an average fit factor improvement of 27.7 for Brand A but only 5 for Brand B. Using tape to seal the edges of the KN95 mask improved fit factor significantly, with an average improvement of 14.7. The use of gauze to seal gaps had a minor improvement of 2.8 while using gauze to bind the mask to the face via the mummy hack improved fit by only 1.6. Tying ear bands resulted in an average improvement of only 0.8.

**Fig. 2:**
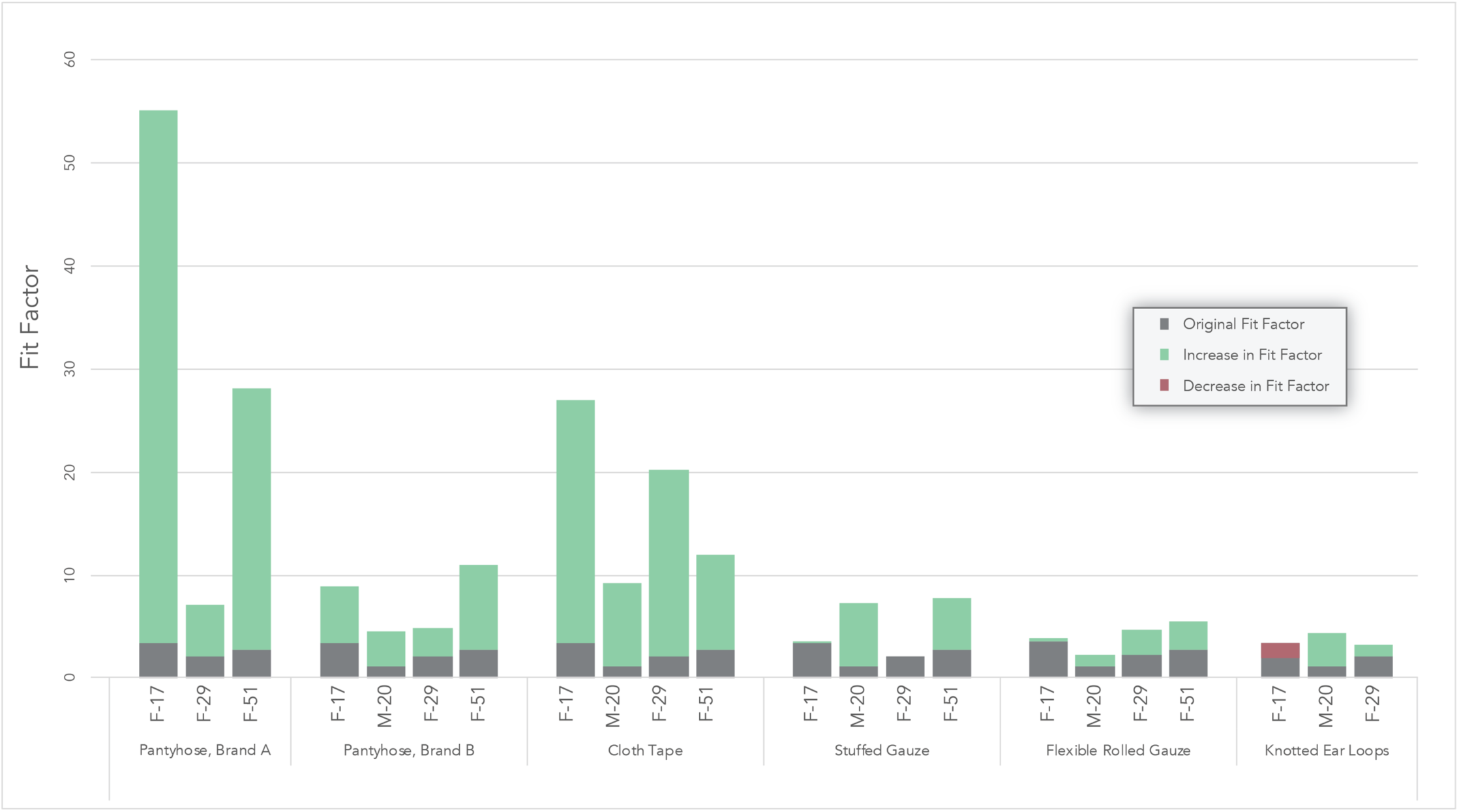
Fit hacks applied to KN95 masks. Gray portions of the bars indicate the performance without the application of a fit hack. Green portions of the bars indicate the amount of improvement. The red portion of the bar indicates the amount of decrease in performance.

Pantyhose and cloth tape improved fit significantly when applied to surgical masks (see Fig. 2). Pantyhose proved effective in improving the fit-factor of surgical masks, with an average improvement of 7.2 for Brand A and 4.9 for Brand B. Tape provided a similar average improvement of 4.8. The least effective hacks were the use of rubber bands, with an average improvement of 2.5, and tying ear bands, with an average improvement of 2.5.

## Discussion

With few exceptions, fit hacks improved the fit factor of both surgical and KN95 masks for all participants. The pantyhose and tape hacks provided most effective for both KN95 and surgical masks, with the rubber band hack showing some promise for surgical masks.

The benefits of each hack differed greatly according to the participant, sometimes in surprising ways. For example, it was expected that fit hacks to make a mask smaller would not significantly benefit participants with large heads. While this proved true for surgical masks, with the individuals with the two smallest heads benefiting the most, it did not hold true for KN95 masks.

An inspection of the fit hacks once applied showed that individual facial features may have a significant impact on fit. For example, a visual inspection of the hacks when applied to participants showed that the nose bridge prevented some hacks from contouring to the sides of the nose, a more significant issue for those with prominent noses (see Fig. 4).

**Fig. 3:**
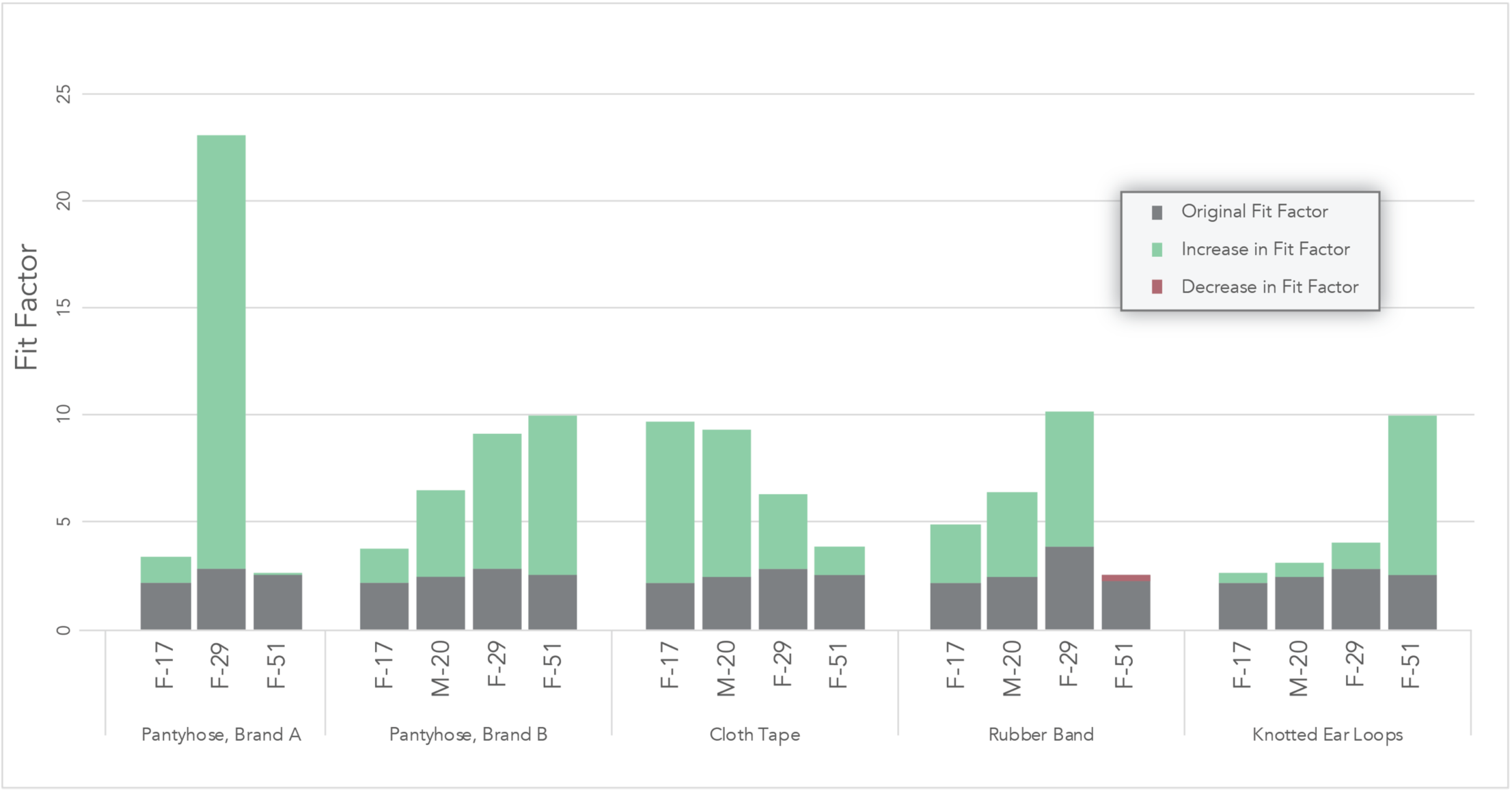
Fit hacks applied to surgical masks. Gray portions of the bars indicate the performance without the application of a fit hack. Green portions of the bars indicate improvement gained with the application of a fit hack. The red portion of the bar indicates a decrease in performance from applying the fit hack.

**Fig. 4:**
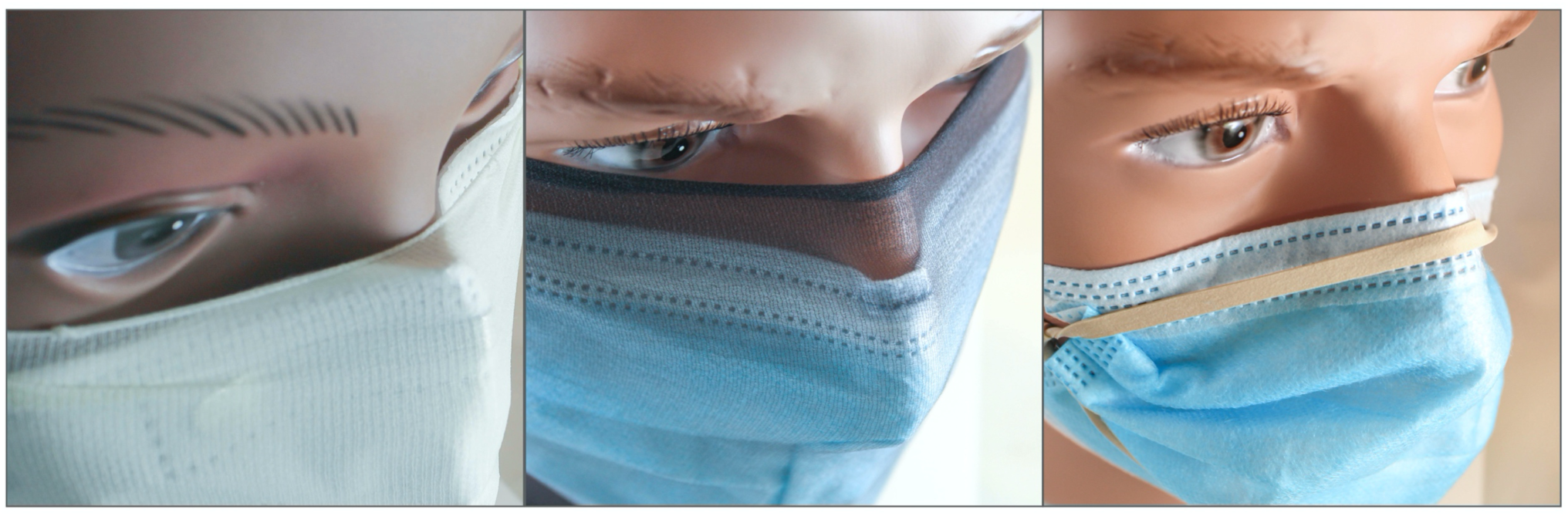
Many fit hacks were unable to fit fix issues around the nose bridge. This impacted wearers with more prominent noses and provided an example of one facial feature critical to the efficacy of fit hacks.

Discomfort was an issue with many of the hacks. The most discomfort was reported with pantyhose and the rubber band hacks. The rubber band hack was found to put extremely painful pressure on the ears and face, going so far as to hinder circulation to the ears for some participants. The pantyhose caused high levels of discomfort as well as issues speaking and occasional obstruction of the eyes. The use of tape was reported as mostly comfortable when worn, but a moderate to high level of discomfort accompanied the tape’s removal.

The most effective and reliable fit hack was the use of pantyhose. Placing pantyhose over the mask and head proved an effective way to improve fit, though it should be noted that the brand of pantyhose used was found to have a significant impact on the benefits incurred. We used large sizes of Brand A and Brand B for our experiment. A section of the thigh was cut to be placed around the head. The material of Brand A was found to be tighter and less flexible. While it did not fit over one participant’s head, it created an improvement which was, on average, approximately two times greater than that of Brand B. The use of pantyhose generated an improvement for all testers, although one tester was unable to fit the pantyhose over their head.

The use of tape to seal the edges of a mask was a similarly reliable hack. Participants who benefited from the hack the most had an assistant to assure the tape was correctly placed and no gaps were present. How cloth tape would seal the mask over longer periods of time is unknown as sweat or movement would be expected to degrade the seal of all but the most flexible tape.

Higher fit scores were achieved when hacks were applied to the KN95 mask. This is likely due to the high filtration but poor fit of the unaltered KN95 mask and indicates that using KN95 masks with fit hacks can potentially provide high levels of protection. One participant was unable to fit the KN95 mask with ear bands tied. Another participant was unable to fit one brand of pantyhose over his head without the pantyhose tearing. In these cases, the problematic hack was not tested.

Although the surgical masks in this study were purchased from a medical supplier and marketed for medical use, there are indications that the masks, made in China, were not certified by the Chinese authorities for medical use. We have learned through discussion with chief administrators at several hospitals that this is a problem facing many hospitals, clinics, and import companies in the United States. It is possible these masks were not medical grade, and that if the hacks were applied to a medical grade mask higher fit scores would be achieved.

## Conclusion

Our results indicate that fit hacks have the potential to significantly increase the fit of face masks, with an improvement of almost 800% having been recorded. Notably, substantial heterogeneity exists regarding the benefit a hack will offer. We identified two key contributory factors: the type of mask and the facial features of the wearer. The most effective hacks tested in this study were the application of tape to seal the edges of the mask and placing pantyhose over the mask. Further research with significant samples sizes is needed to determine the reliability and average improvement of these and other hacks.

## Data Availability

Data used will be freely availible on the Cambridge University open data repository "Apollo".

